# Antenatal Iron-Folic Acid Supplementation Gaps in Nigeria: A Predictive Machine Learning Analysis of Non-Initiation and Sub-Optimal Duration Using NDHS 2023–24

**DOI:** 10.64898/2026.09.14.26363020

**Authors:** Eloghosa Aisosa Nosa-Ihaza, Eze Nnamdi David, Wol Bol Geng Wol, Adeosun Joshua Oluwatimilehin, Uyioghosa Nosayise Nosa-Ihaza

## Abstract

**Background:** Although iron-folic acid (IFA) supplementation is a known, low-cost intervention to prevent anemia in pregnancy, the Nigeria Demographic and Health Survey (NDHS) 2023–24 shows that 33% of women do not begin IFA supplementation, and the majority of women who start IFA supplementation do not complete it for the recommended duration. No nationally representative dataset from Nigeria has formally tested whether these are one or two distinct problems.

**Methods:** This is a cross-sectional secondary analysis of the NDHS Individual Recode - Women aged 15-49 (2023-24). Two gaps were identified: Gap A: Non-initiation of antenatal IFA supplementation (n=13,940); Gap B: Inadequate duration (<90 days) among those who initiated the supplementation (n=8,431). We compared the survey-weighted logistic regression model with elastic net, random forest, and XGBoost models, all of which included the normalised DHS sampling weight, and tested them on an equal 80:20 test-set split held out in Stata. The factors that were found to predict included: education, wealth, residence, geopolitical zone, religion, ethnicity, health insurance, media exposure, access barriers, parity, ANC visit count, timing of the ANC visits, and type of facility. We cross-checked self-reported IFA status with hemoglobin and anemia biomarkers from the same survey.

**Results:** Non-initiation prevalence was 33.8% and among initiators, 59.8% consumed iron for < 90 days. The single factor that dominated Gap A was women with no antenatal contact, who were 21 times more likely not to have initiated than those receiving home-based ANC (95% CI 11.6–38.6), while both public- and private-sector facility care were protective. Education, wealth, and ANC visit count were independently protective for both outcomes. Gap B showed a more diffuse predictor profile and weaker or reversed geographic effects relative to Gap A. XGBoost discrimination was higher in both cases (AUC 0.922 and 0.721) than in survey-weighted logistic regression (AUC 0.915 and 0.678), in contrast to the best comparator study in the literature, which found XGBoost to underperform simpler models for a different health outcome. The directionality of self-reported IFA status relative to the biomarkers (hemoglobin and anemia) was the same for both outcomes (p<0.05).

**Conclusions:** The IFA supplementation shortfall in Nigeria can be attributed to two issues: a large, highly concentrated access gap, primarily driven by the absence of antenatal contact, and a smaller, more diffuse adherence gap not adequately explained by survey variables. Plausibly, two different interventions are needed to close this gap: expanding the reach of antenatal services for non-initiation, and investigating side effects, supply continuity, and household barriers for non-adherence among antenatal service initiators, likely in a qualitative format.

## 1. Introduction

Anemia in pregnancy continues to be one of the most chronic maternal health challenges in low- and middle-income countries. Anemia in pregnancy is estimated at 41.8% of women worldwide, and is significantly higher in sub-Saharan Africa (1), with rates over 55%. Nigeria is particularly impacted by this burden: national survey-based estimates from the 2018 NDHS showed anemia prevalence among pregnant women had reached 61.1% and that there was significant variation by geopolitical zone, with women with less education and lower household wealth consistently having a higher risk of anemia (2,3). Anemia in pregnancy is not only a laboratory diagnosis, but a commonly quoted systematic review and meta-analysis showed that maternal anemia and low antenatal iron consumption are associated with significantly increased low birth weight, preterm delivery, and perinatal mortality (7).

The World Health Organization (WHO) currently recommends that pregnant women take iron and folic acid (IFA) daily during pregnancy as a key intervention to prevent and treat this type of anemia, as part of the basic package of care for pregnant women (8,9). Strong evidence supports this recommendation: a Cochrane systematic review of daily iron supplementation during pregnancy found it 70% more effective at reducing the risk of maternal anemia at term and approximately 57% more effective at reducing iron-deficiency anemia in particular (10). Cost is low, it is readily available in the public health system, and it is already introduced to Nigeria’s antenatal platform. This is a solved problem, in principle. It is not in practice, however: The recently released Nigeria Demographic and Health Survey (NDHS) estimates that only 67% of women with a live birth in the past 12 months supplemented with iron during their last pregnancy, while only a quarter of all women (approximately a third of those who began) supplemented for the recommended duration of 90 days or more (14).

This study focuses on the gap between an intervention’s proven effectiveness and its use in practice across the population. Critically, the NDHS summary statistics already suggest that this is not a homogeneous problem, but two separate and much larger problems: a large proportion of women do not initiate supplementation, and another larger proportion of women who do start do not continue for the recommended duration. This distinction is not made in most of the existing Nigerian literature on IFA supplementation. Studies based on facilities have identified variables such as antenatal counseling, burden of side effects, and health facility attributes as correlates of ‘compliance’ (15, 16), but generally use a small number of facilities in a single geographic location and do not use nationally representative data to measure compliance but rather a single composite outcome. A cross-country analysis of IFA distribution and consumption through antenatal care (17) illustrates the usefulness of using a harmonized, survey-representative approach to answering this question, but Nigeria has not yet experienced a nationally representative survey analysis of IFA distribution and consumption in the country’s newest round of surveys, nor one that specifically addresses the access barrier (does a woman ever begin supplementation?); nor one that addresses the quality/adherence barrier (does she continue it long enough to benefit?).

This distinction matters because the two barriers may stem from different causes and therefore require different policy remedies. Non-initiation is mechanically linked to antenatal care contact: If a woman does not attend ANC and/or attends ANC only sporadically, then she has no opportunity to be offered iron tablets for the first time. Non-adherence among women who do begin, by contrast, is more likely to be attributable to side-effect burden, the continuity of pill supply, or counseling quality, or to other intra-household factors that can affect non-adherence whether or not a woman has reached a facility or not. Most previous studies treat these as a single “compliance” outcome, which may mask the impact of increased access to ANC or enhanced adherence support within the ANC program on the gap for a specific sub-group of women. This study takes a two-stage cascade approach, similar to an access versus quality cascade approach used for antihypertensive treatment gaps, and extended here for the first time to antenatal micronutrient supplementation in Nigeria, which is divided into two gaps, Gap A (not using IFA supplementation) and Gap B (terminating IFA supplementation early).

Another gap in the literature that this study seeks to fill is methodological. Most studies on determinants of IFA adherence in Nigeria and sub-Saharan Africa (15–19) use conventional logistic regression to examine IFA adherence. The relatively few machine learning models identified in this literature, those that are non-linear (e.g., regularised regression, random forests, and gradient boosting) have not been used as extensively as more linear models, and the few instances in which they have been applied to a closely related outcome have yielded mixed results: a 2025 study of predictors of uncontrolled hypertension using the Mexican ENSANUT survey found that, for this particular outcome, XGBoost (a gradient-boosting algorithm) performed worse than simpler random forest and logistic regression models (30), an unexpected finding given that gradient boosting models are generally thought to outperform simpler models. A single study in a different outcome domain, survey, and country context is not enough to determine whether this pattern generalizes to other contexts. This paper directly tests this by fitting four algorithms (survey-weighted logistic regression, elastic net, random forest, and XGBoost) to the same Nigerian data and comparing their out-of-sample discriminative performance.

Lastly, self-reported supplement use may suffer from recall bias, a common problem in almost all adherence studies using surveys (15–17). The NDHS can also cross-check the plausibility of self-reported IFA status with an objective physiological measure (NDHS hemoglobin biomarkers), which earlier research on this cascade could not do because it relied on datasets without comparable biomarker information (32).

This study, therefore, aims to estimate the prevalence of and identify socioeconomic, geographic and health-system predictors of non-initiation of antenatal iron-folic acid supplementation (Gap A) among Nigerian women with a recent live birth, and of sub-optimal duration of IFA supplementation below the 90-day threshold (Gap B) among women who initiated; comparing the predictive performance of survey-weighted logistic regression with three machine-learning algorithms (elastic net, random forest, XGBoost) for both outcomes; and assessing as a partial validation exercise, whether self-reported IFA status is directionally consistent with hemoglobin concentration and anaemia status measured in the same survey round.

## 2. Literature Review

### 2.1 The Burden of Anaemia in Pregnancy and the Case for Iron-Folic Acid Supplementation

Anemia in pregnancy is one of the best documented and most poorly treated maternal health issues worldwide. According to the latest global estimates by the World Health Organization, the prevalence of anemia among pregnant women is around 41.8%, and it is predominantly in sub-Saharan Africa, where over half of pregnant women are affected (1). Nigeria is at the upper end of this distribution. Based on the NDHS 2018, Adeyemi et al. reported national anemia prevalence in pregnancy as 61.1% with significantly high odds of anemia for women in the North Central and South South regions compared with women in the North West, and a distinct education gradient – women with no education had more than double the odds of anemia than women with tertiary education (2). This trend was confirmed using a separate structured additive regression analysis of the same survey round, which included spatial random effects at state level, and which also showed that low education, unemployment, and recent sexually transmitted infection (STI) were risk factors for lower hemoglobin concentration, while states in the South East of the country, especially Imo, had the highest burden nationally (3). The prevalence range of 55 - 75% is consistent across country-level facility-based studies from Rivers State (4), Lagos (5), to a multi-site Lagos/Kano cohort (6), lending credence to the fact that this is not a result of survey methodology but rather a true, geographically widespread characteristic of maternal health in Nigeria.

The clinical consequences of this burden are known. For more than a decade, WHO has included maternal anemia and low maternal iron intake as factors that meaningfully increase the risk of low birth weight, preterm delivery, and perinatal mortality (7), supported by Haider and colleagues’ systematic review and meta-analysis for the Nutrition Impact Model Study Group. The 2012 guideline advocates daily supplementation during pregnancy as standard antenatal care (8), which the 2016 antenatal care guideline reiterated and incorporated (9), and which is WHO’s main response: daily iron and folic acid supplementation. The evidence base for iron supplementation is particularly robust for an intervention to improve public health: a Cochrane systematic review by Peña-Rosas et al. (10) showed a 70% reduction in the risk of maternal anemia at term and a 57% reduction in the risk of iron-deficiency anemia, with iron supplementation carried out daily. IFA supplementation is simple and cheap, requires no specialist skill to administer, and is already part of the nominal package of antenatal care in Nigeria, unlike many maternal health interventions. This study aims to explore not so much whether the intervention is effective, but why it does not work at the population level as intended.

### 2.2 The IFA Supplementation Coverage Gap: Global, Regional, and Nigeria-Specific Evidence

The effectiveness of IFA and its use is well established in low-- and middle-income countries, where reported coverage rates vary widely depending on the setting and how coverage is measured. The variability in reported IFA compliance figures across studies in the systematic review and meta-analysis suggests significant differences in actual IFA compliance rates between countries, as well as methodological differences in how studies defined “compliance” (13). The amount of country-specific evidence is enormous in Ethiopia: a national systematic review and meta-analysis estimated pooled adherence to IFA supplementation at 41.4% (11); and another national meta-analysis estimated pooled adherence at 46.2%, with site-specific estimates ranging from as low as 58.9% in Tigray to as high as 60% in Addis Ababa (12) — which shows how much local health-system and population factors matter, separate from the efficacy of the intervention itself. Similar variability is observed outside Africa – for example, compliance was estimated at 54% in the Philippines and 82% in Brazil, the latter indicating that adherence was poorer with increasing tablet dose frequency, and in Mozambique, adherence to treatment (“two or more ANC visits”) was significantly higher than adherence to treatment (“full course of tablets”) (79% vs. 67%) (17) – an early empirical observation before the present study, that non-initiation and non-completion may be two separate phenomena, with different correlates, rather than a single undifferentiated outcome.

In Nigeria, the most reliable and up-to-date estimates come from the new 2023-24 NDHS. Of women with a live birth in the two years before the survey, 67% used iron supplements during their last pregnancy, of which 25% (about 37%) used it for at least 90 days; a full third of women did not take iron supplements at all (14); the difference between women in rural and urban areas is large, with only 57% of rural women having taken iron compared to 83% of urban women. These figures are similar to, if not better than, earlier facility-based Nigerian estimates: Ugwu and colleagues indicated that the burden of comorbidities was the main driver of poor compliance among antenatal attendees in Enugu (15), while Onyeneho and colleagues’ study across urban, peri-urban and rural communities in the southeast of Nigeria suggested that antenatal counseling exposure and residence type vary in the extent to which they affect uptake of recommended micronutrient amounts needed to prevent anaemia (16). The geographically targeted nature of both studies, typical of Nigeria’s literature on this subject, and the focus on compliance as a single binary or ordinal outcome limit both studies’ generalisability to national policy and their ability to differentiate between access and adherence barriers.

One methodological advance to note is a cross-country study on IFA distribution and consumption via antenatal care (17), which highlights both the feasibility and the usefulness of shifting from single-country, facility-based designs to harmonised, survey-representative designs — though it too treats adherence as a single outcome rather than distinguishing initiation from duration, and Nigeria was not among the countries it examined in depth.

### 2.3 Determinants of IFA Non-Initiation and Non-Adherence

#### 2.3.1 Socioeconomic and Demographic Determinants

Maternal education and household wealth are the most consistent and significant predictors of IFA-related outcomes in almost all studies reviewed and are also the primary predictors of anemia (2,3). Antenatal care attendance and maternal education were the strongest correlates of adherence in Denbiya district (18) and in other Ethiopian facility-based studies (12,19). The association of parity and maternal age is not consistent; some studies report a protective effect of higher parity (which may be interpreted as more experience navigating the health system) while others report no significant association, with education and wealth being the most likely explanation for this apparent association, since the effects of these likely operate through their association with socioeconomic status.

#### 2.3.2 Health System and Antenatal Care Determinants

Antenatal visits, in number and timing, predict IFA outcomes and mechanistically serve as a gatekeeper for iron tablet intake, as they are usually dispensed at ANC visits. The relationship between the use of antenatal care and adherence to iron supplementation (20) is plausible in the context of the study by Karyadi and colleagues in Indonesia, since the factors influencing ANC access also influence IFA outcomes, and education and wealth are known to shape ANC access, which in turn shapes the opportunity to take and be counselled on iron supplementation (21,22). This framing of barriers in terms of supply-chain and health-system factors preceding individual patient behaviour is extended in the cross-country IFA-distribution synthesis (17), which explicitly includes the type and quality of the facility the woman accesses as a barrier as an additional predictor: the type and quality of the facility a woman reaches is plausibly likely to shape whether women are offered supplementation and how.

#### 2.3.3 Geographic, Cultural, and Household-Level Determinants

Nigeria’s well-documented geographic inequality in maternal health service utilisation is not specific to the IFA literature. The utilisation of antenatal care, institutional delivery, and postnatal care services across geopolitical zone, urban/rural residence, and socioeconomic status was jointly explained by significant variation nationwide (23). The findings for ANC visit frequency, using count-data models on previous rounds of NDHS, indicated that the web of geopolitical zone, place of ANC, wealth status, and education explained most of the variation in ANC visits (21), while a study on ANC service utilisation in Jigawa state reinforced the significance of distance, cost, and household decision-making authority as structural barriers unique to the North West region (24). An ethnically oriented study by Fagbamigbe and Idemudia which examined factors affecting utilisation of ANC and found that there was a disparity between the ethnic groups (with higher odds for ANC utilisation among all the major ethnic groups, compared to the Hausa/Fulani, in urban areas while in rural areas, it was not the case in every setting — and in some settings, the Hausa/Fulani had higher odds for ANC utilisation than other ethnic groups), and effects of media on odds of ANC utilisation was found to vary by place of residence, with TV exposure being significant in urban areas, but not in rural areas, and with radio exposure being significant in rural areas, but not in urban areas — with the direction and magnitude of the effects not following a single consistent pattern (24). This previous discovery is directly relevant to, and corroborates, the counterintuitive media-exposure pattern that emerges from models surfaced in this study (television as expected, radio running in the opposite direction).

### 2.4 The Cascade-Of-Care Framework and Its Extension Beyond HIV

The analytic structure employed in this study — dividing a population into those who never initiate an intervention and those who do but fail to adhere (or quality) to it or complete it sufficiently — is most succinctly captured by the HIV “cascade of care” (or “treatment cascade”) framework, which was formally described and quantified by Gardner and colleagues in 2011 (25). The main principle of the cascade framework (i.e., there are categorically different points of failure and categorically different interventions in other chronic disease and preventive care settings) has since been applied to other cascade frameworks. The same principle of access versus quality/adherence has been applied to the present study, as well as the larger program of which it is a part, when considering antenatal micronutrient supplementation, and the two stages of the cascade — non-initiation (Gap A) and sub-optimal duration among initiators (Gap B) — are exactly the same as previously described for chronic disease management. To the best of our knowledge, this is the first study to explicitly report on the two-stage cascade framing, such that “compliance” or “adherence” is not used as a unified outcome rather than a cascade of outcomes, despite the descriptive statistics existing in the literature (as reviewed above) indicating that initiation and completion of IFA supplementation may occur differently, as demonstrated in Mozambique.

### 2.5 Machine Learning Approaches to Predicting Maternal and Reproductive Health Outcomes From DHS Data

In the maternal and reproductive health literature, the use of machine learning methods has increased significantly over the last few years, and the results from these algorithms demonstrated in this large and expanding body of literature are informative for the current study’s comparison of four algorithms. Several studies using DHS data have found that tree-based ensemble models outperform logistic regression on discrimination measures: An application of a random forest model to skilled birth attendance across 27 sub-Saharan African countries found good precision and good accuracy (26); A comparison of four modelling techniques (random forest, XGBoost, logistic regression and decision tree) to predict pregnancy loss in the region of sub-Saharan Africa showed that the random forest model performed best, with an AUC of 94% (27); A study of seven algorithms (random forest, XGBoost, logistic regression, and decision tree, interpreted using SHAP) for health facility delivery in Somalia also preferred the ensemble methods (28). The analysis conducted in Memon and colleagues’ study in Uganda is closest to the present study (29), as they tested logistic regression against six tree-based models (including XGBoost, LightGBM, and CatBoost) for predicting skilled birth attendance, which XGBoost outperformed, with an AUC of 0.75, and found that education, ANC visit count, region, distance to facility, and urban/rural residence are the dominant predictors via SHAP analysis. In this study, XGBoost’s feature-importance analysis for Gap A yielded a remarkably similar profile of predictors, with education, ANC visit count, region, distance to facility, and urban/rural residence as the most significant predictors.

The study reporting predictors of uncontrolled hypertension for 2025 Mexico is a true outlier in the performance of tree-based ensemble models in the DHS literature: logistic regression, LASSO, random forest, and XGBoost were compared, and the latter performed worse than the simpler methods (30). This work differs from the overall trend in the DHS-related literature, where tree-based ensembles are assumed to consistently outperform logistic regression (as much of the maternal-health ML literature would go on to predict) or consistently underperform logistic regression (as the single closest work to this study found). This work fits the four algorithms on the same Nigerian dataset and reports the result as an empirical finding, not an assumed one inherited from either trend. It is also important to note that the inclusion of elastic net, instead of a purely L1-penalised logistic regression, is a small but welcome methodological contribution that stems from the documented superiority of elastic net over other models in dealing with sets of correlated predictor blocks (e.g., wealth and education) typically found in socioeconomic data from the DHS. Most studies in this literature group that include comparative analyses prefer L1-Penalised logistic regression, ordinary logistic regression, or a variety of tree-based models.

### 2.6 Validating Self-Reported Health Behaviour Against Biomarkers

Recall bias and social desirability bias are well documented in self-reported survey measures of health behaviours across a variety of research settings, including this study. The evolving DHS Program biomarker collection efforts are specifically stated to complement self-reported information by providing an objective understanding of health conditions that cannot be fully confirmed through self-report (31). However, despite this stated purpose, few published DHS-based analyses of IFA supplementation have used concurrent hemoglobin biomarker collection to cross-check self-reported supplementation status; thus, this study is a valuable, modest secondary analysis that takes up this methodological opportunity.

### 2.7 Synthesis: Gaps This Study Addresses

That there is a clear body of literature showing that anaemia in pregnancy is a severe problem in Nigeria specifically (2–6) and also in SSA in general; and that, despite the proven low cost high efficacy of IFA supplementation as an intervention, the population-level delivery of this intervention is significantly lower than it could be (7,8,10,14); and that there is a heavy bias in the existing Nigerian and broader SSA evidence base towards facility based, single-outcome studies that do not distinguish access barriers from adherence barriers (13,15,16,18,19); and that attempts to predict maternal health using machine learning methods based on the DHS have gained in popularity over the years, but there is no consensus on which algorithm yields the best results, as the only study directly comparable on the same dataset yielded results contrary to the general trend (XGBoost underperforms) (26–30). To our knowledge, there is no published study or analysis based on the new NDHS 2023-24, which has applied an explicit access/quality cascade framework to the context of IFA supplementation in Nigeria, nor an analysis that has integrated this framework with a rigorous, four-algorithm predictive comparison and a biomarker-based cross-check of self-reported IFA supplementation status. This study addresses all four gaps.

## 3. Methods

### 3.1 Study Design and Data Source

This study is a cross-sectional secondary analysis of the 2023–24 Nigeria Demographic and Health Survey (NDHS), a nationally representative household survey implemented by the National Population Commission and Federal Ministry of Health and Social Welfare with technical support from ICF through the Demographic and Health Surveys (DHS) Program (14). The fieldwork took place between 1st December 2023 and 7th May 2024 in 36 states and the Federal Capital Territory (FCT) with 1,400 Primary Sampling Units (PSUs) and an estimated 42,000 households (14). In the present study, we used an analytic file of individual interviews, as this dataset contains 39,050 women aged 15-49. The sample design, stratification, and weighting follow the principles in the Guide to DHS Statistics (44).

Because the DHS Program obtains ethical clearance for each national survey through the implementing country’s institutional review procedures before data collection and uses only de-identified public-use data, the secondary analysis did not require new ethical approval. DHS Program access to data was provided after normal registration.

### 3.2 Study Population

Because the birth was recent enough and the women were asked the antenatal iron-folic acid (IFA) module, the analytic sample was limited to women. This resulted in an eligible sample of 13,940 women for the primary outcome (Gap A), with 28 women responding “don’t know” to sample question 2 (sample item non-response). After excluding 844 women who answered “don’t know” about the days they supplemented with iron tablets or syrup, the secondary outcome (Gap B) included only a subsample of women who reported using or purchasing iron tablets or iron syrup in their last pregnancy (8,431 women).

### 3.3. Outcome Definitions

#### Gap A — Non-initiation of antenatal IFA supplementation

Coded 1 when the woman had not been given or had not purchased iron tablets or syrup in her last pregnancy; Coded 0 when the woman had been given or had purchased iron tablets or syrup in her last pregnancy.

#### Gap B — Sub-optimal duration of IFA supplementation

For women who started use, coded 1 if she used the supplement for less than 90 days, and 0 if she used it for 90 days or more (Gap A = 0). We used a≥90-day threshold to denote “adequate” supplementation, as this is the threshold commonly used for comparability across surveys of adherence and aligns with the threshold reflected in the NDHS’s published summary tables (14). We note that, although not stated as an ideal duration by WHO, this is a widely used program-monitoring proxy, as opposed to the duration indicated by WHO, which is the entire course of pregnancy, about 180 days; this is discussed further in the Discussion.

### 3.4 Data Quality Screening

We checked all numeric variables for DHS standard “don’t know” and “not applicable” codes before entering them into a model, since these are often included in continuous numeric variables but are actually discrete responses. Two such cases were found and corrected in this screening: the variable representing the number of ANC visits (m14_1) and the variable representing the timing of the first ANC visit (m13_1) are both coded as 98 for “don’t know”, totaling 369 and 22 women, respectively, and both were recoded to missing before being used as continuous predictors. It is this step that deserves special description since it was not detected during routine checking alone: an initial specification containing the uncorrected m14_1 resulted in a pseudolikelihood value that did not change after more than 1400 iterations, which is diagnostic of a pathological outlier (a few women with an implausible value of 98 ANC visits) and not of natural model complexity or of computational limitation. After correction, the equivalent full model converged in less than 4 seconds. We report this as a methodological note, as the same uncorrected variable had been used in preliminary machine-learning models (random forest, XGBoost) without causing loud failure events, and only by comparing results before and after this correction was a failure event detected. It is suggested that all continuous DHS recode variables be systematically checked for embedded sentinel codes as a routine practice, as a non-converging maximum-likelihood estimator may not indicate this type of problem, penalised, or ensemble methods will not necessarily alert to such an issue.

More screening led to two additional specification decisions. First, raw ethnicity (v131) has many distinct groups, with most having fewer than 20 women, so entering it directly produced dozens of values for quasi-complete separation and absurdly large effect estimates (odds ratios greater than 400 with equally huge confidence intervals). Before using it in any model, we reduced it to the four largest nationalities (Hausa, Yoruba, Igbo, and Fulani) and the “Other/Minority” category. Second, in the Gap B analytic sample, the category of religion “other” (v130, code 96) had only 3 female respondents and yielded a small sample coefficient for a zero-width confidence interval, so these women (16 in the full dataset) were recoded as missing on this variable instead of keeping them in a specification that would have led to a false impression of precision.

### 3.5 Predictor Variables

There were nine controls in each of these models: highest level of education (no formal education, primary education, secondary education, higher education); household wealth quintile; type of place of residence (urban, rural); geopolitical zone; religion; ethnic group (categorised as above); health insurance coverage; frequency of reading a newspaper (ordinal), listening to radio (ordinal), and watching television (ordinal); DHS “problems in accessing health care” battery (permission to travel alone, money for treatment, distance to facility, each a binary “problems big” indicator). Maternal age, parity (total number of children ever born), and number and timing of antenatal visits during the last pregnancy were identified as study-specific predictors. We created a single-category facility-type variable with four categories (home, public-sector facility, private-sector facility, or no ANC attendance), from the multiple-response place-of-ANC items in the DHS by prioritizing the formal-facility category over home-based care when women reported more than one place of ANC attendance during their pregnancy for Gap A.

### 3.6 Statistical Analysis

#### Descriptive statistics

Survey-weighted percentages were calculated for categorical variables, and weighted means with standard deviations were calculated for continuous variables and summarised by the outcome status.

#### Survey-weighted logistic regression

We corrected singletons and used the normalised individual sampling weight (NISW) in place of common primary sampling units (PSUs) for clustering and stratification; we fitted survey-weighted logistic regression for each outcome in Stata 15.1 (StataCorp, College Station, TX) using Taylor-series linearisation. Because the outcome samples are limited subpopulations of the full women’s sample, we used the subpop() option instead of restricting the sample first to the outcomes and then to the women, which would have adversely affected the full survey design used for variance estimation (44). To serve as an independent cross-validation, the same model was estimated independently in Python 3.12 with generalized estimating equations (GEE) with an independence working correlation structure and a cluster-robust variance estimator clustered on the primary sampling unit (33) and using the normalised sampling weight as a case weight, through the Python statsmodels package. The point estimates from both implementations were similar throughout (e.g., odds ratio for ANC visit count and for timing of first ANC visit matched to three decimal places between the two implementations and the general property that the stratification term in the Stata linearisation is not included in the GEE approach is expected to give slightly wider confidence intervals than the fully design-based approach).

#### Machine-learning comparison

Three additional algorithms were fitted to each outcome: elastic net regularised logistic regression (34), random forest (35), and extreme gradient boosting (XGBoost) (36), fitted using Python scikit-learn and XGboost libraries, respectively, with normalised sampling weight passed as the out-of-the-box sample_weight argument of each estimator. The 80/20 train/test partition was defined once in Stata, with a fixed random seed applied separately (and independently) to the Gap A and Gap B eligible samples, and exported as a case-level flag to allow each algorithm (including the logistic comparator) to be tested against the same women who were not used for training. The partition was confirmed to be balanced on outcome prevalence on the train and test sets prior to use. Random forest models were fit with 500 trees and a max tree depth of 6 and a minimum leaf size of 20; XGBoost models were fit with 300 boosting rounds, max tree depth 4, learning rate 0.05, and 80% row/column subsampling; and an elastic net used an L1:L2 mixing parameter of 0.5. In this analysis, the results are comparative and exploratory; therefore, the hyperparameters were fixed a priori. This four-algorithm design is directly based on the closest methodological comparison found in the literature, an analysis of predictors of uncontrolled hypertension conducted with the methodology of XGBoost and three simpler algorithms in Mexico in 2025 (30), which was also conducted in this study, but with a different outcome, survey, and country context.

#### Biomarker cross-validation

Weighted mean hemoglobin concentration (altitude-adjusted, g/dL) and prevalence of any anemia were compared by weighted Welch’s t-tests within the women who had valid hemoglobin measurements, between women from the two Gaps (Gap A and Gap B). This is directionally suggestive, not causal evidence, because hemoglobin was measured at the same interview as the retrospective IFA report, and reverse causal pathways (anaemic women being offered iron preferentially) cannot be ruled out.

We used the normalised weight of individual women from the DHS sample for all analyses, and set the significance level at α=0.05. All analyses were performed in Stata 15.1 and Python 3.12 (pandas, statsmodels, scikit-learn, and XGboost).

## 4. Results

### 4.1 Sample Characteristics and Analytic Flow

The total number of women aged 15–49 years interviewed in the 2023–24 NDHS was 39,050; 13,940 had a valid (not “don’t know”) response to the question on antenatal iron-folic acid (IFA) supplementation, constituting the Gap A analytic sample. Of these, 9,275 (66.2% unweighted, 66.2% weighted) said they were given or bought iron tablets/syrup during their last pregnancy, and 4,665 (33.5% unweighted, 33.8% weighted) reported they were not given or bought iron tablets/syrup during the last pregnancy (near the national level of 33% non-initiation reported in the NDHS published summary). Of the 9275 women who started supplementation, 844 answered “don’t know” about the duration and were excluded, leaving a total of 8431 women in the Gap B analytic sample: 3453 women (40.9%) took iron for 90 days or more and 4978 (59.1% unweighted, 59.8% weighted) took iron for less than 90 days.

**Figure 1.**
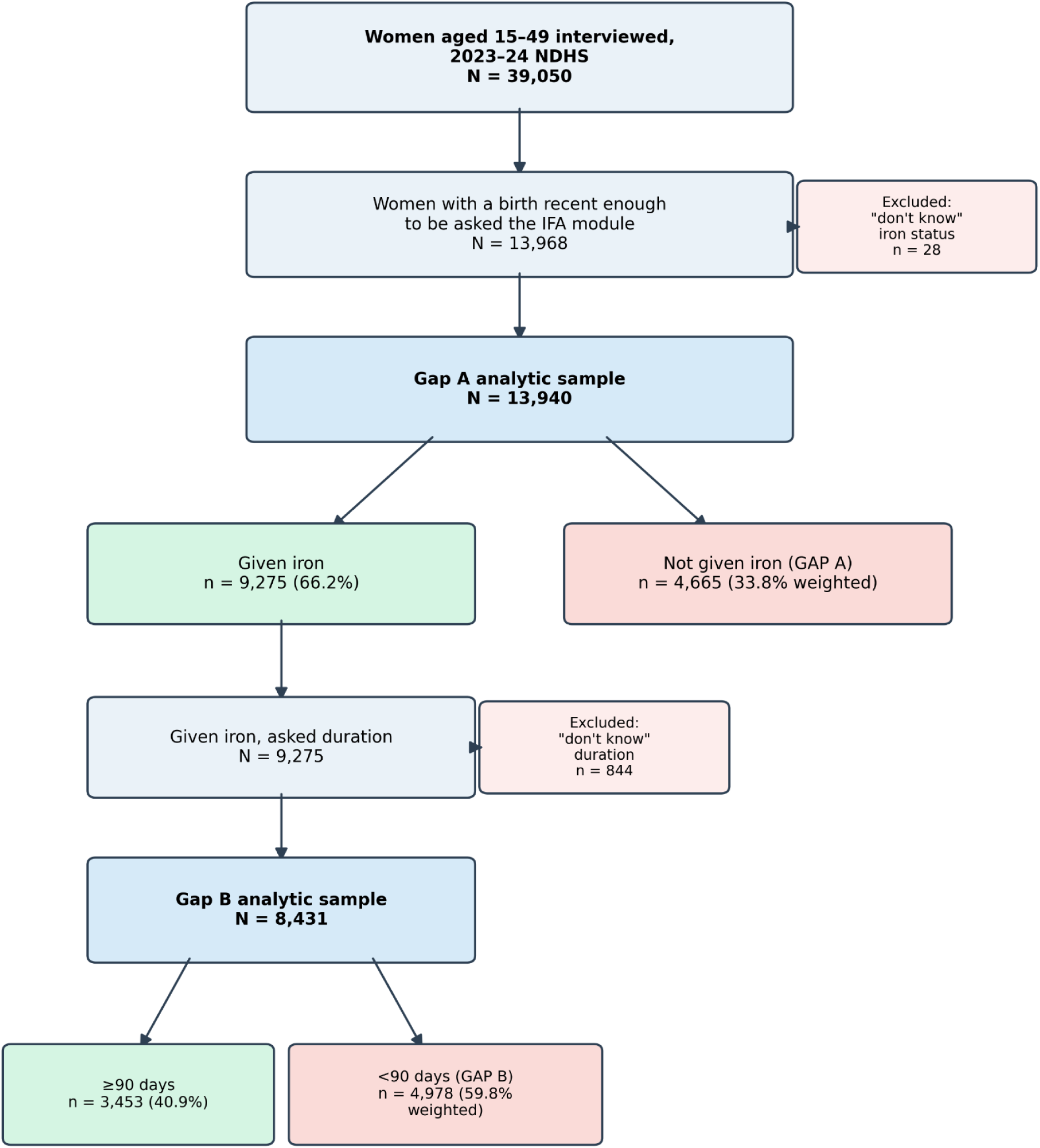
Sample selection flow diagram for the Gap A and Gap B analytic samples.

**Table 1.** Selected weighted characteristics by Gap A status.

| **Characteristic** | **Given iron (n=9,275)** | **Not given iron (n=4,665)** |
| --- | --- | --- |
| Education: No education | 33.3% | 72.2% |
| Education: Primary | 12.4% | 9.6% |
| Education: Secondary | 40.4% | 15.8% |
| Education: Higher | 13.8% | 2.5% |
| Wealth: Poorest | 16.5% | 40.0% |
| Wealth: Poorer | 19.2% | 29.7% |
| Wealth: Middle | 21.2% | 17.5% |
| Wealth: Richer | 22.4% | 8.6% |
| Wealth: Richest | 20.8% | 4.3% |
| Residence: Urban | 47.5% | 19.9% |
| Residence: Rural | 52.5% | 80.1% |
| Zone: North West | 34.7% | 48.8% |
| Zone: North East | 22.0% | 16.2% |
| Zone: North Central | 13.2% | 22.8% |
| Zone: South East | 7.8% | 3.7% |
| Zone: South South | 9.6% | 3.7% |
| Zone: South West | 12.8% | 4.9% |
| "Permission to go alone" a big problem | 9.1% | 19.0% |
| Age, mean (SD) | 29.4 (6.7) | 28.7 (7.2) |
| Parity, mean (SD) | 3.7 (2.5) | 4.0 (2.5) |
| ANC visits, mean (SD) | 5.90 (4.09) | 1.28 (2.80) |

The contrast of these figures by each socio-economic and access dimension is stark and consistent across the board: women not given iron were more than twice as likely to have no education (72.2% vs. 33.3%), nearly four times more likely to be in the poorest wealth quintile (40.0% vs. 16.5%), overwhelmingly rural (80.1% vs. 52.5%), and attended on average fewer than one-quarter as many antenatal care visits (1.28 vs. 5.90).

### 4.2 Gap A: Predictors of Non-Initiation of IFA Supplementation

The survey-weighted logistic regression model (Stata svy: logit, subpopulation of 13,564 women after excluding item non-response across all covariates; design-based F(36,1271)=64.58, p<0.001) showed that the following were significant predictors of non-initiation:

**Table 2.** Survey-weighted logistic regression, Gap A (non-initiation)

| **Predictor** | **OR** | **95% CI** | **p** |
| --- | --- | --- | --- |
| Education: Primary (ref: none) | 0.60 | 0.46–0.78 | <0.001 |
| Education: Secondary | 0.52 | 0.41–0.66 | <0.001 |
| Education: Higher | 0.40 | 0.27–0.59 | <0.001 |
| Wealth: Poorer (ref: poorest) | 0.80 | 0.65–0.99 | 0.044 |
| Wealth: Middle | 0.63 | 0.47–0.83 | 0.001 |
| Wealth: Richer | 0.57 | 0.41–0.78 | 0.001 |
| Wealth: Richest | 0.59 | 0.40–0.88 | 0.010 |
| Zone: North East (ref: NW) | 0.72 | 0.55–0.95 | 0.022 |
| Zone: North Central | 2.59 | 1.87–3.59 | <0.001 |
| Zone: South East | 2.43 | 1.26–4.68 | 0.008 |
| Zone: South South | 1.56 | 0.99–2.45 | 0.054 |
| Zone: South West | 1.45 | 0.88–2.40 | 0.147 |
| Facility: No ANC (ref: Home) | 21.13 | 11.55–38.64 | <0.001 |
| Facility: Private | 0.34 | 0.19–0.60 | <0.001 |
| Facility: Public | 0.33 | 0.19–0.59 | <0.001 |
| Ethnicity: Yoruba (ref: Hausa) | 2.00 | 1.30–3.07 | 0.002 |
| Religion: Islam (ref: Catholic) | 1.51 | 1.04–2.17 | 0.029 |
| Radio <1/week (ref: $\geq 1/\text{wk}$) | 0.68 | 0.52–0.90 | 0.006 |
| Radio: not at all | 0.65 | 0.52–0.81 | <0.001 |
| TV: not at all | 1.32 | 1.00–1.74 | 0.046 |
| Parity | 0.93 | 0.88–0.97 | 0.001 |
| ANC visit count | 0.95 | 0.92–0.98 | 0.003 |

Urban/rural residence, health insurance, newspaper exposure, and the money/distance access-barrier items were not significant once the other covariates were accounted for. Facility type had the biggest effect in the model: women who reported no antenatal contacts had 21 times the odds of not initiating care compared with those who had home-based care—a greater difference than any socioeconomic predictor. There was no meaningful public/private gradient once a woman reached any formal facility (public vs private facility care: 0.33 vs. 0.34).

### 4.3 Gap B: Predictors of Sub-Optimal Duration Among Women Who Initiated

In the equivalent analysis of Gap B (subpopulation n=8,100; design-based F(34,1273)=15.00, p<0.001), the pattern, in general less strong, was the opposite.

**Table 3.** Survey-weighted logistic regression, Gap B (sub-optimal duration)

| Predictor | OR | 95% CI | p |
| --- | --- | --- | --- |
| Education: Primary (ref: none) | 0.67 | 0.55–0.81 | <0.001 |
| Education: Secondary | 0.68 | 0.56–0.82 | <0.001 |
| Education: Higher | 0.57 | 0.44–0.74 | <0.001 |
| Wealth: Poorer (ref: poorest) | 0.78 | 0.63–0.96 | 0.020 |
| Wealth: Middle | 0.68 | 0.54–0.85 | 0.001 |
| Wealth: Richer | 0.75 | 0.57–0.98 | 0.032 |
| Wealth: Richest | 0.61 | 0.46–0.82 | 0.001 |
| Zone: North Central (ref: NW) | 1.36 | 1.05–1.76 | 0.020 |
| Zone: South South | 1.51 | 1.10–2.06 | 0.010 |
| Zone: South West | 2.30 | 1.53–3.44 | <0.001 |
| Ethnicity: Other/Minority (ref: Hausa) | 0.75 | 0.58–0.95 | 0.019 |
| Permission not a big problem (ref: is) | 0.51 | 0.40–0.66 | <0.001 |
| Parity | 0.96 | 0.92–0.99 | 0.023 |
| Timing of 1st ANC visit (per month later) | 1.16 | 1.11–1.22 | <0.001 |
| ANC visit count | 0.88 | 0.86–0.90 | <0.001 |

The relationship of education and wealth to Gap B was similar to Gap A, though the geographic effects reversed or weakened: Some of the same areas that had an effect on Gap A had a nonexistent or inverse relationship to Gap B, and where zone was significant (North Central, South South, South West), women in those zones had higher odds of being under 90 days, the opposite direction from several of the same zones’ effect on Gap A. Religion, radio/TV exposure, and health insurance were not significant for Gap B. Later initiation of ANC (each additional month before first visit) raised the odds of sub-optimal duration by 16%, which is mechanically sensible, since starting later leaves less remaining pregnancy in which to accumulate 90 days.

### 4.4 Comparison of Predictive Algorithms

The four algorithms were tested against a common, Stata-defined held-out test partition (Gap A: 2,692 test women; Gap B: 1,643 test women):

**Table 4.** Out-of-sample discrimination (AUC) by algorithm.

| Algorithm | Gap A AUC | Gap B AUC |
| --- | --- | --- |
| Survey-weighted logistic | 0.9148 | 0.6777 |
| Elastic Net | 0.9150 | 0.6780 |
| Random Forest | 0.9143 | 0.7023 |
| XGBoost | 0.9218 | 0.7208 |

**Figure 2.**
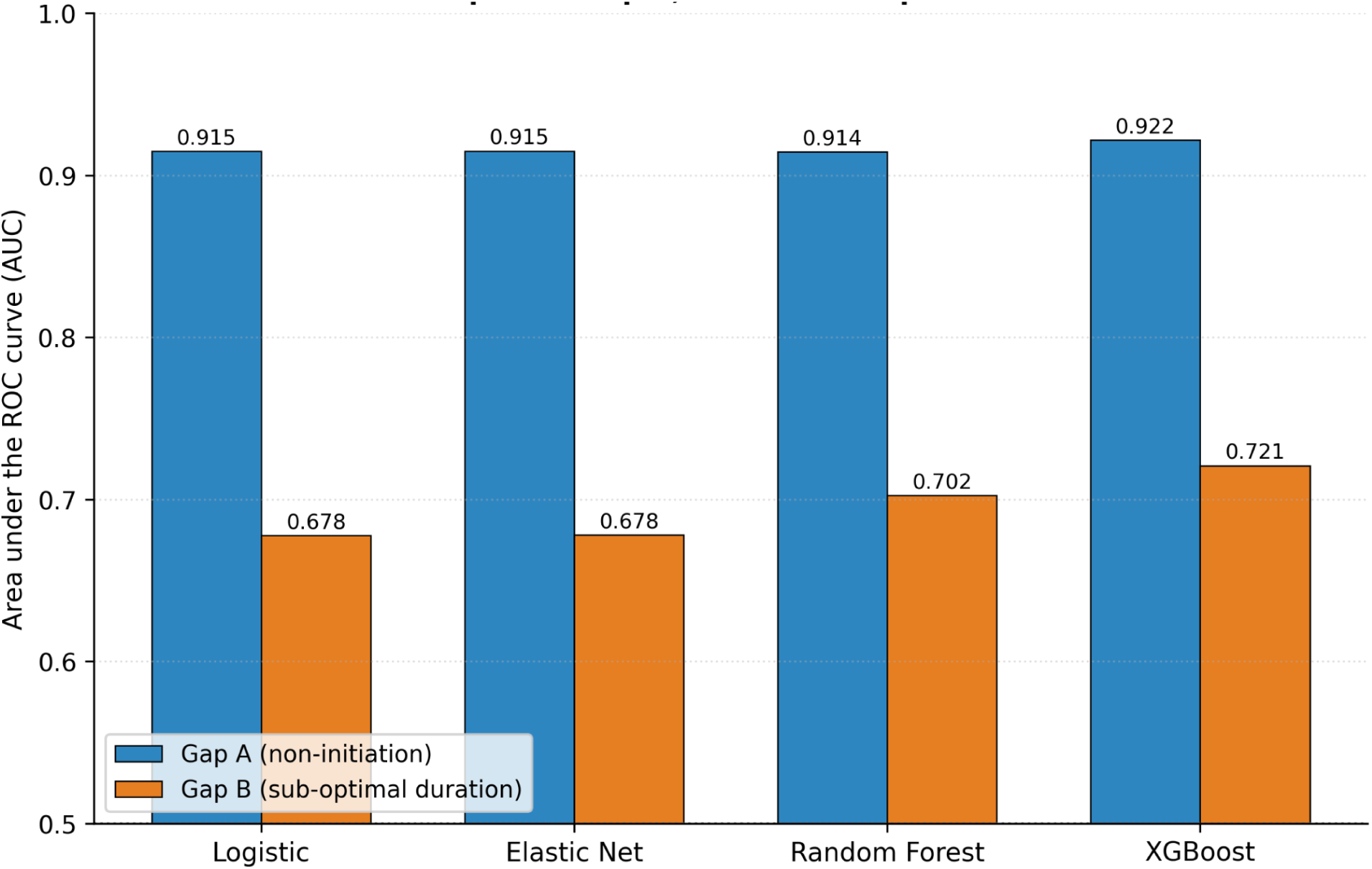
Out-of-sample discrimination (AUC) by algorithm, Gap A vs. Gap B, held-out test partition.

The highest discrimination was obtained by XGBoost on both outcomes, albeit with a relatively small difference to logistic regression for Gap A (+0.007) and a larger difference for Gap B (+0.043). This is the reverse of the closest study in the literature (Mendoza-Cano et al., ENSANUT Mexico, hypertension), which saw XGBoost outperform simpler methods. The improvement with elastic net over unpenalised logistic regression was negligible for both outcomes (+0.0002 and +0.0003), suggesting little redundant or correlated structure among predictors to regularise. Across algorithms, Gap A was significantly more predictable than Gap B (AUC range 0.914-0.922 vs. 0.678-0.721), reflecting the robust and coherent predictor signal for Gap A (combined facility type and ANC visit count).

This asymmetry is even more emphasised by the feature importance of the final XGBoost model. No other single predictor accounted for more than 2% of total model importance; for Gap A, lack of any antenatal contact made up 57%, while ANC visit count was a distant second (12%). Importance for Gap B was much more spread out: ANC visit count (6.8%), no education (5.0%), timing of the first ANC visit (3.7%), and the permission-to-travel barrier (3.6% and 2.5% across the two categories). There was no single dominant driver.

**Figure 3.**
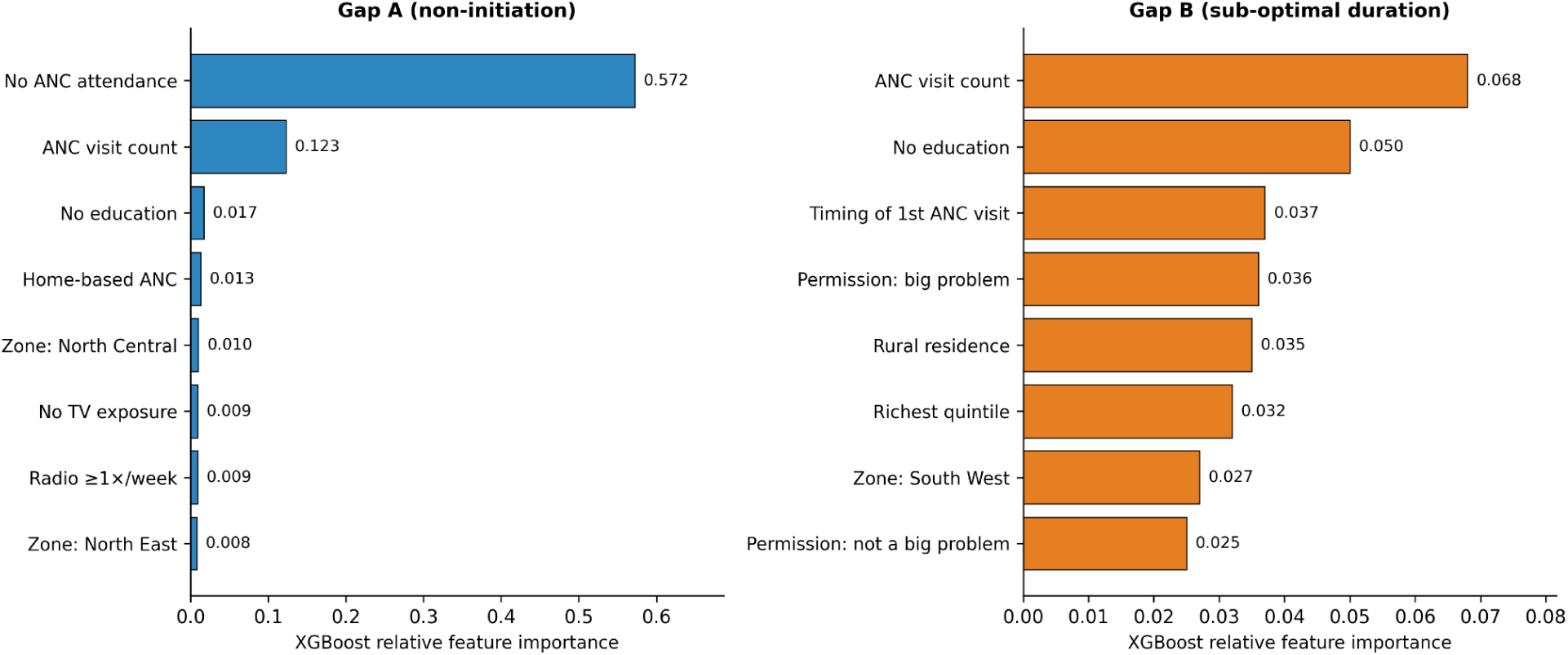
Top 8 XGBoost feature importances, Gap A and Gap B.

### 4.5 Biomarker Cross-Validation

Among women with a valid hemoglobin measurement, self-reported IFA and objectively measured hemoglobin and anemia status showed consistent directionality for both outcomes.

Women given iron had higher mean hemoglobin than women not given iron (12.07 g/dL, SD 1.48, n=3,839 vs. 11.90 g/dL, SD 1.60, n=1,755; weighted Welch’s t=-3.96, p<0.001) and lower prevalence of any anemia (40.4% vs. 44.7%). Among women who initiated supplementation, those who sustained it for 90 days or more had higher hemoglobin than those who fell short (12.13 g/dL, SD 1.41, n=1,420 vs. 12.02 g/dL, SD 1.54, n=2,013; t=-2.27, p=0.023) and lower anemia prevalence (38.0% vs. 41.7%). Both associations are statistically significant but small in magnitude and, as noted in the Methods, provide directionally supportive evidence of the self-report validity of the two measures, not causal evidence, because both were measured at the same time, in the same interview.

## 5. Discussion

### 5.1 Principal Findings

This study revealed that IFA supplementation shortfall in Nigeria is two-fold and has very different profiles. Whether a woman had any antenatal contacts (Gap A, 33.8% weighted prevalence) is the dominant factor in non-initiation. Women with zero ANC attendance had 21 times the odds of non-initiation compared to women who attended even the home-based ANC, and type of ANC facility accounted for 57% of the XGBoost model’s total predictive weight, far outweighing the combined effect of education, wealth, and geography. Among women who initiated, sub-optimal duration was a much more complex pattern; no one factor dominated, geographic associations were not as strong or as positive as those for Gap A, and predictive discrimination was significantly lower for each algorithm employed (AUC 0.68–0.72 compared with 0.91–0.92 for Gap A). This asymmetry is not a constraint; it is a key result: it shows empirically that there are two different mechanisms underlying access and adherence to IFA supplementation, and thus two different policy levers – the organising principle of the overall research program this study is part of.

### 5.2 Interpreting Gap A: An Access Problem, Not a Supply Problem

The size of the “no ANC” effect on Gap A should be thought about, as it changes the policy challenge. The protective effect of public- and private-sector facility care was nearly the same (OR 0.33 vs. 0.34); once a woman reaches some formal point of antenatal contact, the type of facility she reaches seems to matter little in determining her likelihood of receiving iron. The binding constraint is reaching a facility at all—consistent with the ANC coverage gap found nationwide in Nigeria, where national-level antenatal attendance has been reported to feature high geographic variation; wealth, education, and geopolitical zone explain most differences in ANC visits. The higher odds of non-initiation in North Central and South East than in the North West region, where raw ANC coverage estimates tend to be the lowest in the country, is counterintuitive and should be treated with caution, but it is not a simplistic explanation. The model may already adjust for facility type and the number of ANC visits, which could explain the zone effect. This is a hypothesis the present data cannot adjudicate, and it is a logical topic for further qualitative or health-systems follow-up studies, but not for further secondary DHS analysis.

Since 2005 and updated regularly (most recently in 2021), Nigeria has National Guidelines for the Prevention and Control of Micronutrient Deficiencies that specifically recommend daily IFA during pregnancy and include formulation options aimed at minimising side effects and enhancing uptake (37). Given this 21-fold access gap (mostly based on whether women access antenatal care) and that this is a national guideline rather than a local one, the binding constraint on access is not the guideline’s content but reach and coordination. A 2019 assessment of Nigeria’s micronutrient deficiency control programming found insufficient coordination across government, private-sector, and civil-society interventions, and flagged the persistent difficulty of reaching target populations as a central risk to program effectiveness (38) — a diagnosis this study’s facility-type finding corroborates from the demand side, more than a decade after the guideline’s introduction.

### 5.3 Interpreting Gap B: Adherence as a Distinct, Harder Problem

This may be due to the fact that several factors considered as being key to adherence, such as side effects of the pills, perceived size or taste of the pill, continuity of supply at the facility level, and quality of counseling at initiation, were not accounted for in the DHS instrument, leading to a less pronounced predictor profile for Gap B and significantly lower predictability than for Gap A. This analysis is limited to what DHS collects, but Nigerian studies in Enugu and southeastern Nigeria showed side effects and counseling exposure as determinants of adherence, which Nigeria’s national guideline explicitly names as adherence levers. This is consistent with, and helps explain, why the ceiling of each algorithm was so much lower in Gap B than in Gap A: the best and most powerful predictors of adherence might not exist in this dataset. It is a significant exception and one that might make a difference: the permission to travel alone barrier was not just Gap A — it was Gap B (OR 0.51), whether or not she actually made the trip, or whether she was able to return to replenish supplies or attend follow-up counseling throughout the pregnancy.

### 5.4 The Algorithm Comparison in Context

XGBoost performed better than the survey-weighted logistic regression on both outcomes, in direct opposition to the closest comparator study in this literature, ENSANUT Mexico hypertension analysis in 2025, where XGBoost performed worse than simpler models (30). It’s not that gradient boosting is generically better; it’s that it’s more likely to be better in certain circumstances: For Gap A (+0.007 AUC over logistic), the single-predictor structure is fairly dominant (facility type), which is exactly the kind of signal gradient boosting can’t do much better than a logistic regression on the single predictor, and for Gap B (+0.043 AUC over logistic), the more diffuse, potentially non-linear or interactive structure of the predictors is exactly the kind of signal gradient boosting can do better than logistic regression. This is in line with the general methodological finding that flexible algorithms outperform linear models more when true relationships are non-additive, and underperform or gain little benefit in the presence of a strong signal concentrated in a few strong predictors, which the ENSANUT comparator study’s single hypertension outcome could not, by design, demonstrate. The minimal improvements in AUC by adding the predictor set used in this study to the “standard” predictor set (wealth and education) for regularisation (by elastic net) compared to pure LASSO suggests that this study’s specific predictor set has low multicollinearity, which is the block where regularisation is expected to offer additional benefit.

### 5.5 Biomarker Cross-Validation: A Partial, Not Definitive, Reassurance

However, the essentially consistent, statistically significant, and directionally correct association between self-reported IFA and both hemoglobin concentration and anemia prevalence, with both IFA gaps, suggests that self-reported IFA is not as far from physiological reality as some observers suggest. However, the association is weak in magnitude (a 0.11–0.17 g/dL difference in mean hemoglobin) and reverse causation (because anaemic women were more likely to be offered iron, a potential reason for the association, but in the opposite direction to the true supplementation efficacy) cannot be excluded by the same-interview, cross-sectional study design. This should be interpreted as proof that self-report isn’t typically meaningless, not as proof that there is no recall bias.

### 5.6 Broader Context and Forward-Looking Implications

Nigeria’s pattern — a persistent gap between documented guidelines and delivered coverage, and between initiation and sustained adherence — is not unique to Nigeria. This study’s key message is that interventions to boost IFA supplementation coverage are more likely to be effective when customized to address the specific mechanism driving a subgroup’s failure than when applied in a generic “improve compliance” manner. A recent methodology and state-ranking analysis of the Indian Anemia Mukt Bharat program showed that after years of sustained national investment, IFA supplementation coverage was not the same across states (39), which again supports this study’s point. Going forward, WHO now recommends that Multiple Micronutrient Supplementation (MMS) is generally more effective than IFA alone in preventing low birth weight and small-for-gestational-age birth (40,41), and there is increasing evidence that the effectiveness of MMS is indeed significantly different depending on the level of adherence (42) — likely leaving the access/adherence dichotomy from IFA to MMS unchanged despite a change in product. In 2021, the National Guidelines for the Prevention and Control of Micronutrient Deficiencies formally approved MMS distribution at all ANC and PNC clinics nationally, replacing IFA (37); but the implementation of MMS in Bayelsa State was found to be inconsistent and incomplete in the field during the assessment in 2025, with significant gaps in health-worker training and in the consistency and quality of supply of MMS in local government areas (LGA) (43). For IFA, the access/adherence separation illustrated here is likely to carry over to other products distributed in Nigeria as it moves towards MMS: whatever is causing non-initiation (facility contact) will not go away, and what is causing non-adherence (diffuse, likely side-effect- and counseling-related factors) will not either, as long as the product in question is IFA.

### 5.7 Strengths

The study’s methodological elements are its main advantages. It is based on the recently published, nationally representative, large (n=13,940-39,050 depending on the subpopulation) 2023-24 NDHS, as opposed to the prevailing mode in the existing literature in Nigeria, which was a facility-based convenience sample. To our knowledge, it is the first application of an explicit access/quality cascade framework to IFA supplementation in Nigeria and makes the first head-to-head comparison of all four algorithms (survey-weighted logistic regression, elastic net, random forest and XGBoost) directly and empirically against each other, directly testing whether the ENSANUT comparator study result holds for all of the outcome domains within IFA supplementation. Point estimates were calculated independently in two statistical implementations (Stata’s svy: logit and a Python cluster-robust estimator using GEE) and were generally close across both implementations; many were identical, with some agreeing to three decimal places. A systematic screening process uncovered and rectified a data-coding artifact (a “don’t know” sentinel value in two continuous ANC variables) that would have remained silent in the broader DHS-based analytic literature were it not for the systematic screening process. Lastly, this study also includes a partial validation of the self-reported supplementation status using a biomarker, which has been used to a lesser extent in the published IFA literature and was possible thanks to the concurrent collection of hemoglobin in the DHS.

### 5.8 Limitations

However, some restrictions need to be expressly stated. Firstly, this is a cross-sectional analysis and cannot provide any evidence of causality; any associations mentioned should only be interpreted in predictive or descriptive terms, not in causal terms. Second, maternal self-reported supplementation status and duration are also subject to recall bias, and the biomarker cross-check provides partial (not definitive) reassurance, as explained above. Third, the 90-day cut-off, which is employed as a widely used program-monitoring proxy for adequate supplementation, does not represent WHO’s stated ideal of the full course of supplementation (approximately 180 days), and results using this cut-off should be interpreted as indicative of “adequate supplementation” in the WHO sense rather than “full compliance”. The fourth table shows that only 3.2% of the sample had any type of health insurance, and the analysis is too small to provide enough power to conclude that there was no effect of insurance status on either outcome, but rather that the result was not significant; the absence of significance should be interpreted as not a lack of effect but rather as inconclusive. Fifth, the findings on media-exposure should be interpreted with care, as previous studies of the effects of exposure to the Nigerian ANC similarly have found effects that are not easily summarised in a simple story: television exposure had the desired effect (protective) while radio exposure had the opposite (harmful); the effects on other media, such as newspapers, were not significant. Residual confounding between radio ownership and unmeasured household or rural-infrastructure characteristics, not captured by the study’s media-exposure controls, cannot be ruled out. Sixth, the facility type variable was constructed from a multiple-response question about the entire pregnancy and may not fully reflect a woman’s “usual” or main site of antenatal services when she reported more than one location. Lastly, the hyperparameters for the machine-learning algorithms were set a priori for this study’s comparative purposes rather than performance-maximising objectives, potentially understating the predictive upper limit of each algorithm, especially XGBoost and random forest. Last but not least, these findings are specific to the Nigerian context and should not be applied to other national contexts without independent testing.

## 6. Conclusion

When examined closely, empirically, Nigeria’s iron-folic acid supplementation shortfall during pregnancy uncovers two distinct problems – one an access gap which is large and sharply concentrated and associated almost exclusively with antenatal care contact – and a smaller and more diffuse adherence gap, which survey evidence available suggests is only partly understood. Machine Learning, specifically XGBoost, provided a modest, but real, predictive benefit to logistic regression used in the traditional survey model for both outcomes, particularly the more challenging, more ‘diffuse’ adherence outcome — as this one existing comparator study in a different health domain did not find, and as the ability of flexible algorithms to outperform linear models appears to be outcome-specific and not universal. Inequitable access to IFA will likely need to be addressed through two separate interventions: increasing the reach of antenatal care and timely contact to increase the likelihood of initiation, and a more detailed, perhaps qualitative, examination of side effects, interruptions to IFA supplies, and household-level barriers of non-adherence among the majority of IFA initiators who do not use IFA for the recommended duration.

## Declarations

### Ethics Approval and Consent to Participate

The secondary data used for analysis in this study are de-identified and publicly available from the 2023–24 Nigeria Demographic and Health Survey (NDHS). The original survey team obtained informed consent from all interviewees, and the original NDHS protocol was approved by the National Health Research Ethics Committee (NHREC), Nigeria, and the ICF Institutional Review Board (IRB). This analysis relied on de-identified secondary data that is accessible via registered, authorized access to the DHS Program repository, so it was not reviewed for additional institutional review.

### Clinical Trial Registration

N/A

### Artificial Intelligence Statement

This work is not generated by a Generative Artificial Intelligence (G.A.I.) or large language model (L.L.M.) tool. The information provided is the authors’ own views and opinions.

## Acknowledgements

The authors are thankful to Ahead Labs, IIT Roorkee, for providing Stata 15 and the necessary skillset for this project. The authors also acknowledge Marwadi University for providing the resources that helped in the successful conduct of the research.

## Data Availability Statement

Data analysed in this study are available from the DHS Program (https://dhsprogram.com) but are not publicly available because of restrictions to protect the respondents’ confidentiality. Access is only granted with registration and approval by the DHS Program, upon reasonable request at https://dhsprogram.com/data/available-datasets.cfm. The dataset used was the Individual Recode (women’s) file of the 2023–24 Nigeria Demographic and Health Survey (NDHS). We abstracted additional literature and information from publicly available clinical trial information and World Health Organization (WHO) reports.

